# Ground level utility of AWaRe Classification: Insights from a Tertiary Care Center In North India

**DOI:** 10.1101/2023.08.02.23293536

**Authors:** Gunjita Negi, Arjun KB, Prasan Kumar Panda

## Abstract

**Background:** The overuse and misuse of antimicrobials contribute significantly to antimicrobial resistance (AMR), which is a global public health concern. India has particularly high rates of antimicrobial resistance, posing a threat to effective treatment. The WHO AWaRe classification system was introduced to address this issue and guide appropriate antibiotic prescribing. However, there is a lack of studies examining the prescribing patterns of antimicrobials using the AWaRe classification, especially in North India. Therefore, this study aimed to assess the prescribing patterns of antimicrobials using the WHO AWaRe classification in a tertiary care centre in North India.

**Aim:** To study the prescribing patterns of antimicrobials using WHO AWaRe classification through a cross-sectional study in AIIMS Rishikesh.

**Methods:** A descriptive, cross-sectional study was conducted from July 2022 to August 2022 at a tertiary care hospital. Prescriptions containing at least one antimicrobial were included in the study. Data on prescriptions, including patient demographics, departments, types of antimicrobials prescribed, and duration of treatment, were collected. A questionnaire-based survey was also conducted to assess the knowledge and practices of prescribing doctors regarding the utility of AWaRe classification.

**Results:** A total of 123 patients were included in the study, with antibiotic prescriptions being written for all of them. Most prescriptions were for inpatients, evenly distributed between Medicine and Surgical departments. Metronidazole and Ceftriaxone were the most prescribed antibiotics. According to the AWaRe classification, 57.61% of antibiotics fell under the Access category, 38.27% in Watch, and 4.11% in Reserve. The majority of Access antibiotics were prescribed in the Medicine department, while Watch antibiotics were more common in the Medicine department as well. The questionnaire survey showed that only a third of participants were aware of the AWaRe classification, and there was a lack of knowledge regarding antimicrobial resistance and the potential impact of AWaRe usage.

**Conclusion:** This study highlights the need for better antimicrobial prescribing practices and increased awareness of the WHO AWaRe classification and antimicrobial resistance (AMR) among healthcare professionals. The findings indicate a high proportion of prescriptions falling under the Access category, suggesting appropriate antibiotic selection. However, there is a significant difference between the WHO DDD and the prescribed daily dose in the analysed prescriptions suggesting overuse and underuse of antibiotics. There is room for improvement and educational interventions and antimicrobial stewardship programs should be implemented to enhance knowledge and adherence to guidelines, ultimately contributing to the containment of antimicrobial resistance.

## INTRODUCTION

The discovery of antimicrobials is regarded as one of the most crucial advancements of the 20th century, contributing significantly to saving lives. The appropriate prescription of antimicrobials is essential, focusing on situations where they can provide substantial benefits to patients with optimal effectiveness while minimizing the risk of antimicrobial resistance (AMR) exacerbation(1) However, there is mounting evidence indicating that the overuse and misuse of antimicrobials have become leading factors in the complex causative web of AMR (2) Antimicrobial resistance poses a global public health problem, capable of spreading and causing significant human and economic burdens. The situation in India is particularly concerning, as the country experiences some of the highest rates of antimicrobial resistance among bacteria commonly causing infections in both community and healthcare settings (3).

It is imperative to address the emergence of multi-drug resistant organisms, which threaten the progress made in medical advancements over the past century. Conversely, in many parts of the world, there is not only overuse and misuse of antimicrobials but also inadequate access. Pneumonia remains a leading cause of childhood deaths globally, with over 2 million estimated deaths per year, primarily due to limited access to antimicrobials (4). Thus, promoting access to these life-saving medicines for those in need should also be a priority (1).

Antimicrobial stewardship aims to bridge the gap between excessive and inadequate antimicrobial use (5). However, assessing the impact of antimicrobial stewardship programs (ASPs) has proven challenging, as the commonly used metrics, such as defined daily doses (DDD) per 1000 inhabitants per day and days of therapy, provide limited information about the quality of antibiotic use (6). The existing defined daily dose method used in adult antibiotic surveillance is unsuitable for neonates and children due to their widely variable bodyweights (7).

To address these issues, the World Health Organization (WHO) updated its Model List of Essential Medicines in 2017, introducing the Access, Watch, Reserve (AWaRe) classification system (8). The AWaRe system categorizes antimicrobials based on their appropriateness, safety, and potential impact on AMR. The Access group includes antibiotics of choice for the 25 most common infections, with favourable safety profiles and a low likelihood of exacerbating AMR. These antimicrobials should be consistently available, affordable, and quality-assured. The Watch group comprises critically important antimicrobials that require strict monitoring and limited use due to their higher potential for negatively impacting AMR. The Reserve group includes last-resort antimicrobials effective against multi-drug resistant bacteria, and their use should be minimized as they represent a valuable, non-renewable resource. The fourth category is of the Discouraged antimicrobials, mostly including antimicrobial combinations this was developed in the 2019 EML update (1). The AWaRe system is also represented as a traffic-light approach: Access = green, Watch = orange and Reserve = red. The overall goal was to reduce the use of Watch Group and Reserve Group, and to increase the use of Access antibiotics to >60% by 2023 (4).

Unlike previous measures of antibiotic consumption, the AWaRe classification allows for the quantification of antibiotic use in each category, providing insights into the overall quality of antibiotic use within a country (7). Therefore, the WHO AWaRe categories can serve as a valuable tool for monitoring antibiotic consumption and optimizing antibiotic use, complementing antibiotic stewardship efforts at the national level.

Given the ongoing struggle to identify appropriate measures for hospitals and their antimicrobial stewardship programs, this study aims to audit antimicrobial prescriptions and compare the utility of the AWaRe classification with other process measurements of antimicrobial utilization. Additionally, the study seeks to assess the knowledge and attitudes of prescribing physicians regarding antimicrobial resistance and the AWaRe classification. This study is particularly important in the context of North India, where the burden of resistance is increasing, and limited research has been conducted in this region. Furthermore, the study aims to examine the prescribing practices of antibiotics in a tertiary care center and compare them with the WHO Defined Daily Dosage to determine whether common antibiotics are being under or over-dosed.

### Objectives

1. To audit the prescribing patterns of antimicrobials among clinicians using WHO’s AWaRe classification in a tertiary care centre
2. To assess knowledge and practices of prescribing doctor about the utility of AWaRe by Questionnaire based assessment
3. To compare the appropriateness of AWaRe classification with days of therapy and defined daily doses of antimicrobial utilization

## METHODS

### Study design

Cross sectional in nature

### Setting

This study was carried out in a tertiary care centre in North India. The sample collection was carried out for a duration of 2 months.

### Study population

Physicians (faculty, junior and senior residents, interns) and their prescriptions having antimicrobials

### Inclusion criteria

- Prescriptions containing at least one antimicrobial in both outpatient and inpatient setting.
- Same prescribed physicians

### Exclusion criteria

- Treating physician not giving consent

This is a descriptive, cross-sectional study conducted on patient prescriptions of various departments in a tertiary care hospital from July 2022 to August 2022. The study was conducted after approval by the Institutional Ethics Committee (All India Institute of Medical Sciences, Rishikesh, India(Reference number -AIIMS/IEC/22/252))Prescriptions containing at least one antimicrobial, prescribed to patients of all ages, and in various departments was included in the study after ruling our exclusion criteria.

Universal sampling was used to select prescriptions. The aim of the study was explained clearly to the prescribing doctor and an informed consent form was obtained from willing doctors to fill questionnaires about their knowledge and practices on AWaRe classification. Data on prescriptions, including patient demographics, departments, types of antimicrobials prescribed, and duration of treatment, were collected. The relevance of antimicrobial prescriptions was checked by measuring appropriateness (right drug, right dose and right duration) according to WHO guidelines, Infectious Disease Society of America (IDSA) guidelines and disease specific guidelines if needed.

Following this, the practising physicians’ knowledge and practices towards AWaRe classification was evaluated using a questionnaire-based study by online or offline modes. The questionnaire was self-structured after searching medical literature for comparable studies and adapting questions designed in other physicians’ surveys previously carried out. The questionnaire comprised of two set of questions one for the assessment on of knowledge and the other part for the attitude of the physicians about antimicrobial resistance and AWaRe classification. Pre-validation of questionnaire for its contents and relevance is done by experts (one clinician, one pharmacologist, one biostatistician).

## Statistical analysis

Data was collected on proforma, google form, and excel sheet and analysed by estimating the proportion of various variables including antimicrobial utilisations as per AWaRe classification. Univariate analysis was performed and the data was arranged as percentages of total. Mean daily dosing for a particular antimicrobial was determined and compared with WHO DDD using the Students T test.

## RESULTS

As depicted in Table 1., in this study, a total of 123 patients were included, and each of them received antibiotic prescriptions. Most of the prescriptions were written for inpatients (75.4%) and both the Medicine and Surgical departments were represented equally with 49.6% and 50.4% respectively. Among the healthcare professionals who prescribed antibiotics, 72% were Junior Residents, 18.7% were Senior Residents, and 9.3% were Consultants. The prescriptions included 27 different antibiotics, with Metronidazole being the most prescribed (19%) followed by Ceftriaxone (17%). The WHO AWaRe classification system was used to analyse the antibiotics prescribed, with 57.61% falling under the Access category, 38.27% in Watch, and 4.11% in Reserve Category. The mean number of antibiotics used per patient was 1.84 ± 0.83. The mean duration of antibiotics prescribed was 6.63 ± 3.83 days. The minimum number of antibiotics prescribed per patient was 1, while the maximum was 5.

**Table 1.**
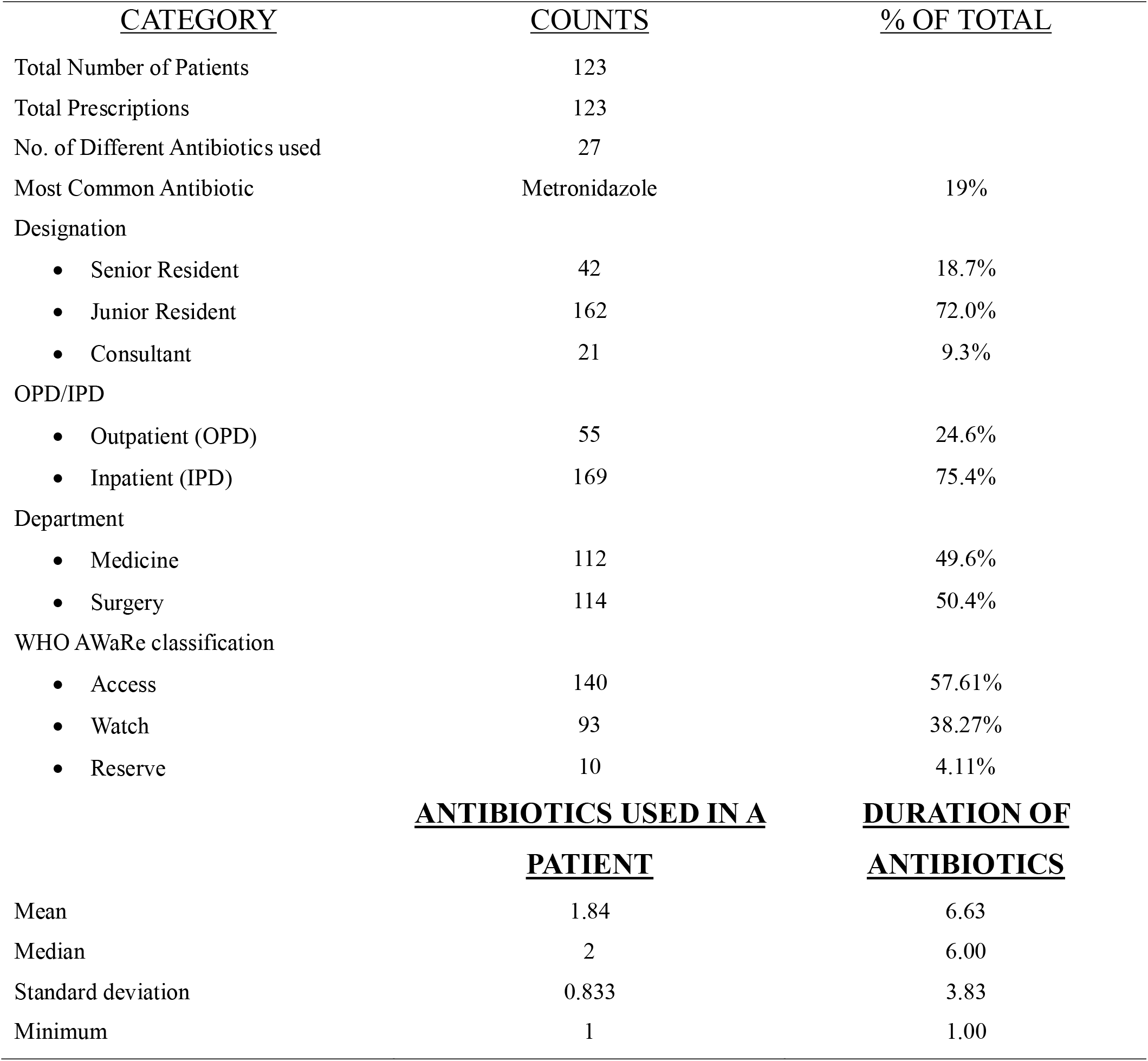
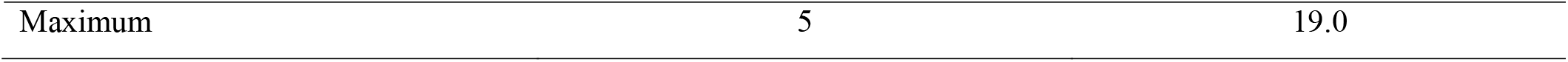
Descriptive data representing the Antibiotic Usage patterns categorized according to the Designation of prescriber, Department, Outpatient vs Inpatient and AWaRe classification. Second half of the table represents the Descriptive analysis of antibiotics used and duration of individual antibiotics.

The distribution of antibiotics prescribed according to the WHO AWaRe categories is presented in Fig 2. The difference in prescribing frequencies amongst departments and can be noted. Among the prescribed antibiotics, 57.61% were categorized as Access, 38.27% as Watch and 4.11% as Reserve. Most of the antibiotics prescribed in the Access category were from the Medicine department (75.4%), followed by Surgery (24.6%). For Watch antibiotics, Medicine had a higher proportion (63.4%) compared to Surgery (36.6%). In terms of seniority, Junior Residents prescribed the highest number of antibiotics for both Access and Watch categories in Medicine and Surgery departments. Senior residents and Consultants prescribed a lower number of antibiotics in all categories and departments. Only a few antibiotics were prescribed in the Reserve category, with most prescriptions being from the Medicine department.

**Fig 1.**
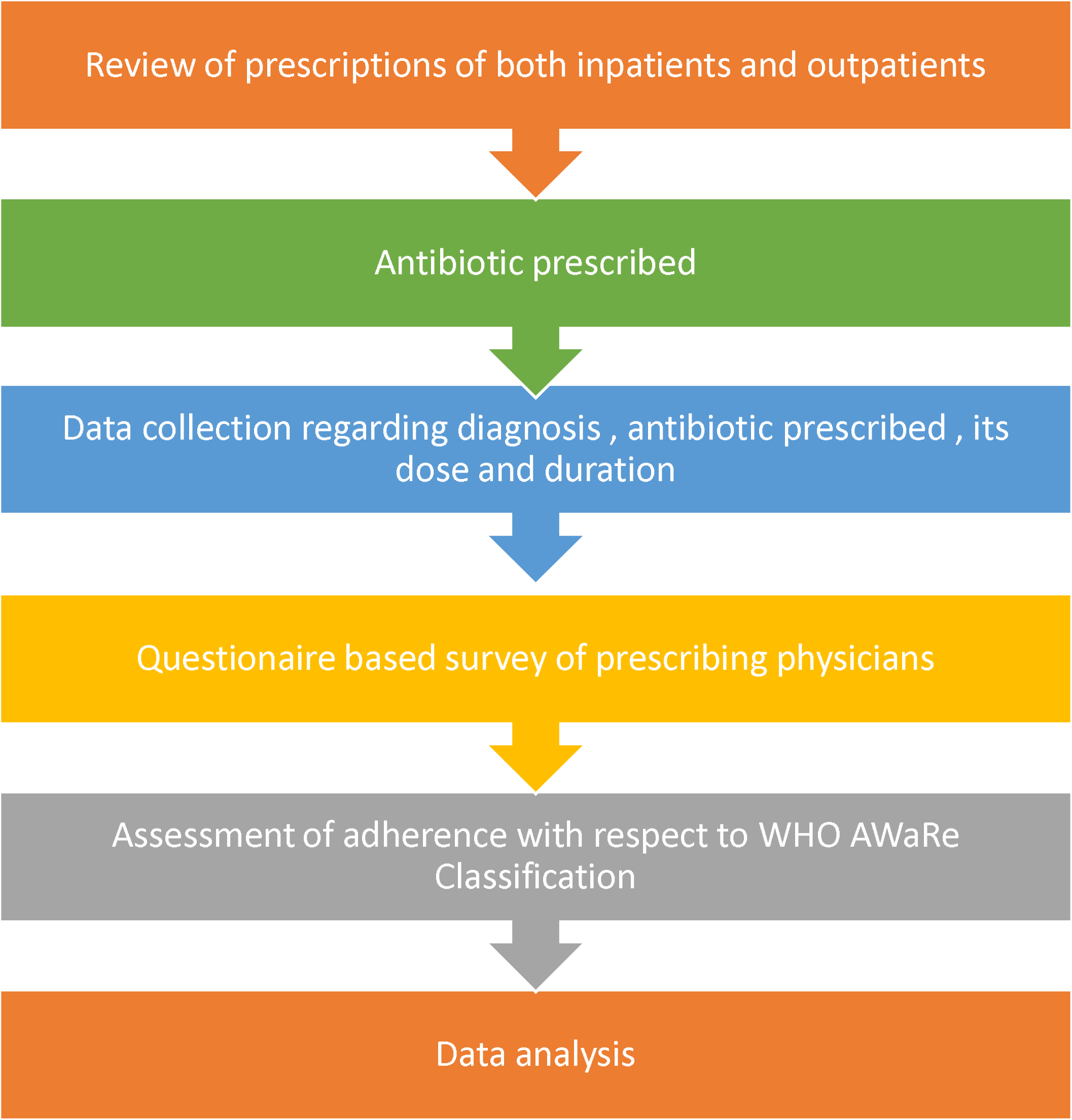
Representation of the study flow

**Fig 2.**
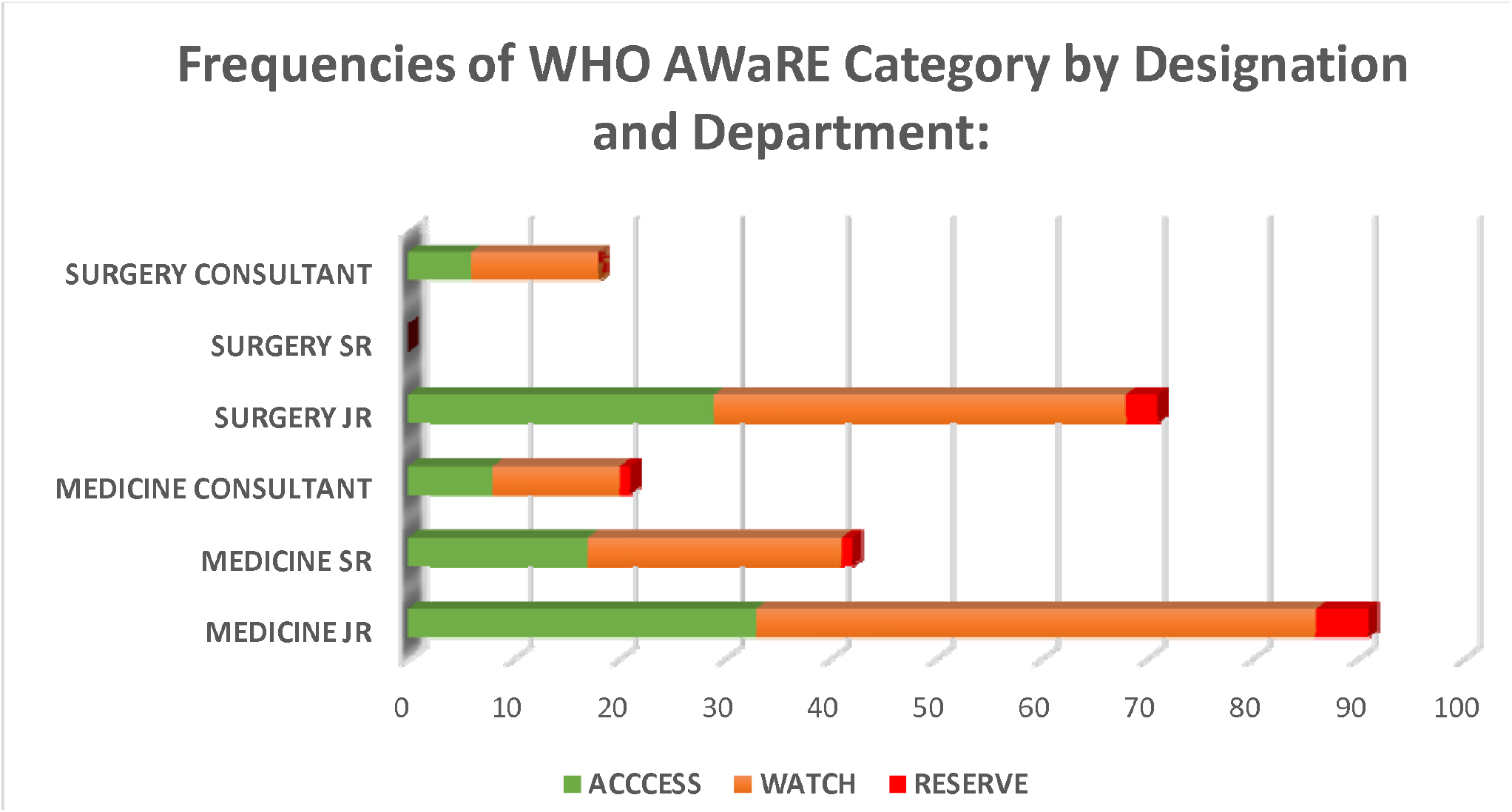
Frequencies of WHO AWaRE Category by Designation and Department

The study also evaluated the Knowledge and Awareness of Healthcare professionals towards the WHO AWaRe classification through a questionnaire survey. A total of 93 participants responded to the survey. Among them, most participants were Junior Residents (69.9%), followed by Senior Residents (25.8%) and Faculty (4.3%). When enquired if they knew about the WHO AWaRe classification only 33.3% of the participants responded positively. Of those who were aware of the AWaRe classification, the most common source of information was the internet (31.2%), followed by the antimicrobial policy of their institution (15.1%).

The survey results on the awareness of antimicrobial resistance among healthcare professionals are also presented in the table 2. Out of the 93 participants, 68 (73.1%) agreed that the emergence of antimicrobial resistance is inevitable, while only 13 (14.0%) disagreed that AWaRe usage will result in the inability to treat serious infections. Additionally, 58 (62.4%) agreed that it will lead to lengthier hospital stays, 43 (46.2%) agreed that the success of chemotherapy and major surgery will be hampered, and the majority also agreed that Its use will lead to increased cost of treatment and increased mortality rates. Regarding the utilization of AWaRe in the hospital summarized in Table 2, 35.5% of the participants agreed that it should be used, while only 2.2% disagreed. Additionally, 34.4% agreed that AWaRe reduces adverse effects of inappropriate prescription. However, 37.6% of the participants considered that AWaRe threatens a clinician’s autonomy and 30.1% thought that its use can delay treatment.

**Table 2.** Representation of Knowledge of WHO AWaRe Classification Among Healthcare Professionals.

| <u>CATEGORY</u> |  | <u>COUNTS</u> | <u>% OF TOTAL</u> |  |  |  |
| --- | --- | --- | --- | --- | --- | --- |
| Position |  |  |  |  |  |  |
| • | Junior Resident | 65 | 69.9% |  |  |  |
| • | Senior Resident | 24 | 25.8% |  |  |  |
| • | Faculty | 4 | 4.3% |  |  |  |
| Do You Know about WHO AWaRe classification |  |  |  |  |  |  |
| • | No Details | 22 | 23.7% |  |  |  |
| • | Yes | 31 | 33.3% |  |  |  |
| • | Never heard | 21 | 22.6% |  |  |  |
| • | Little Idea | 19 | 20.4% |  |  |  |
| How did you hear about AWaRe? |  |  |  |  |  |  |
| • | The internet | 29 | 31.2% |  |  |  |
| • | The WHO website | 10 | 10.8% |  |  |  |
| • | The antimicrobial policy of our institution | 14 | 15.1% |  |  |  |
| • | Other sources | 19 | 20.4% |  |  |  |
| • | No idea about it | 21 | 22.6% |  |  |  |
| <u>Knowledge (Score) on WHO AWaRe Classification</u> |  |  |  |  |  |  |
|  | <u>TOTAL</u><br><u>SCORE</u> | <u>JUNIOR</u><br><u>RESIDENT</u><br>(N=21) | <u>SENIOR</u><br><u>RESIDENT</u><br>(N=9) | <u>CONSULTANT</u><br>(N=3) | <u>MEDICINE</u><br>(N=24) | <u>SURGERY</u><br>(N=9) |
| N | 33 | - | - | - | - | - |
| MEAN | 3.91 | 3.81 | 4.22 | 3.67 | 3.79 | 4.22 |
| STANDARD<br>DEVIATION | 2.17 | 2.25 | 2.33 | 1.53 | 2.08 | 2.49 |
| MINIMUM | 1 | 1 | 1 | 2 | 1 | 1 |
| MAXIMUM | 8 | 8 | 8 | 5 | 8 | 8 |

**Table 3.** Representation of awareness of WHO AWaRe Classification Among Healthcare.

| **<u>Awareness of WHO AWaRe Classification Among Healthcare professionals :</u>** |  |  |  |  |  |
| --- | --- | --- | --- | --- | --- |
| **<u>STATEMENT</u>** | **<u>FALSE</u>** | **<u>TRUE</u>** | **<u>NO IDEA</u>** | **<u>N</u>** |  |
| **Emergence of Antimicrobial Resistance is inevitable** | 24 | 68 | 1 | 93 |  |
| **It will result in inability to treat serious infections** | 13 | 80 | 0 | 93 |  |
| **Lengthier hospital stays will be a result** | 35 | 58 | 0 | 93 |  |
| **Success of chemotherapy and major surgery will be hampered** | 50 | 43 | 0 | 93 |  |
| **Cost of treatment will be increased** | 52 | 41 | 0 | 93 |  |
| **MMR and IMR will increase** | 62 | 31 | 0 | 93 |  |
| **<u>QUESTION</u>** | **<u>STRONGLY DISAGREE</u>** | **<u>DISAGREE</u>** | **<u>NEUTRAL</u>** | **<u>AGREE</u>** | **<u>STRONGLY AGREE</u>** |
| **Should AWaRe be used in the hospital?** | 18 (19.4%) | 2 (2.2%) | 21 (22.6%) | 33 (35.5%) | 19 (20.4%) |
| **AWaRe reduces adverse effects of inappropriate prescription** | 12 (12.9%) | 5 (5.4%) | 25 (26.9%) | 32 (34.4%) | 19 (20.4%) |
| **AWaRe threatens a clinician's autonomy** | 9 (9.7%) | 35 (37.6%) | 35 (37.6%) | 11 (11.8%) | 3 (3.2%) |
| **It can delay treatment** | 16 (17.2%) | 37 (39.8%) | 28 (30.1%) | 8 (8.6%) | 4 (4.3%) |

Additionally, the Defined Daily Dosage of each drug was also evaluated. The usage of various antimicrobial drugs in a hospital setting, along with their daily doses and defined daily dosages (DDD) according to the WHO’s Anatomical Therapeutic Chemical (ATC) classification system was calculated. Some of the important findings include high usage rates of ceftriaxone and metronidazole, and relatively low usage rates of drugs like colistin and clindamycin. Additionally, some drugs had wider ranges than others. Comparison of WHO defined DDD with Daily Drug dose (Mean) in the studied prescriptions is represented in the Clustered Bar chart in Fig 3.

**Fig 3.**
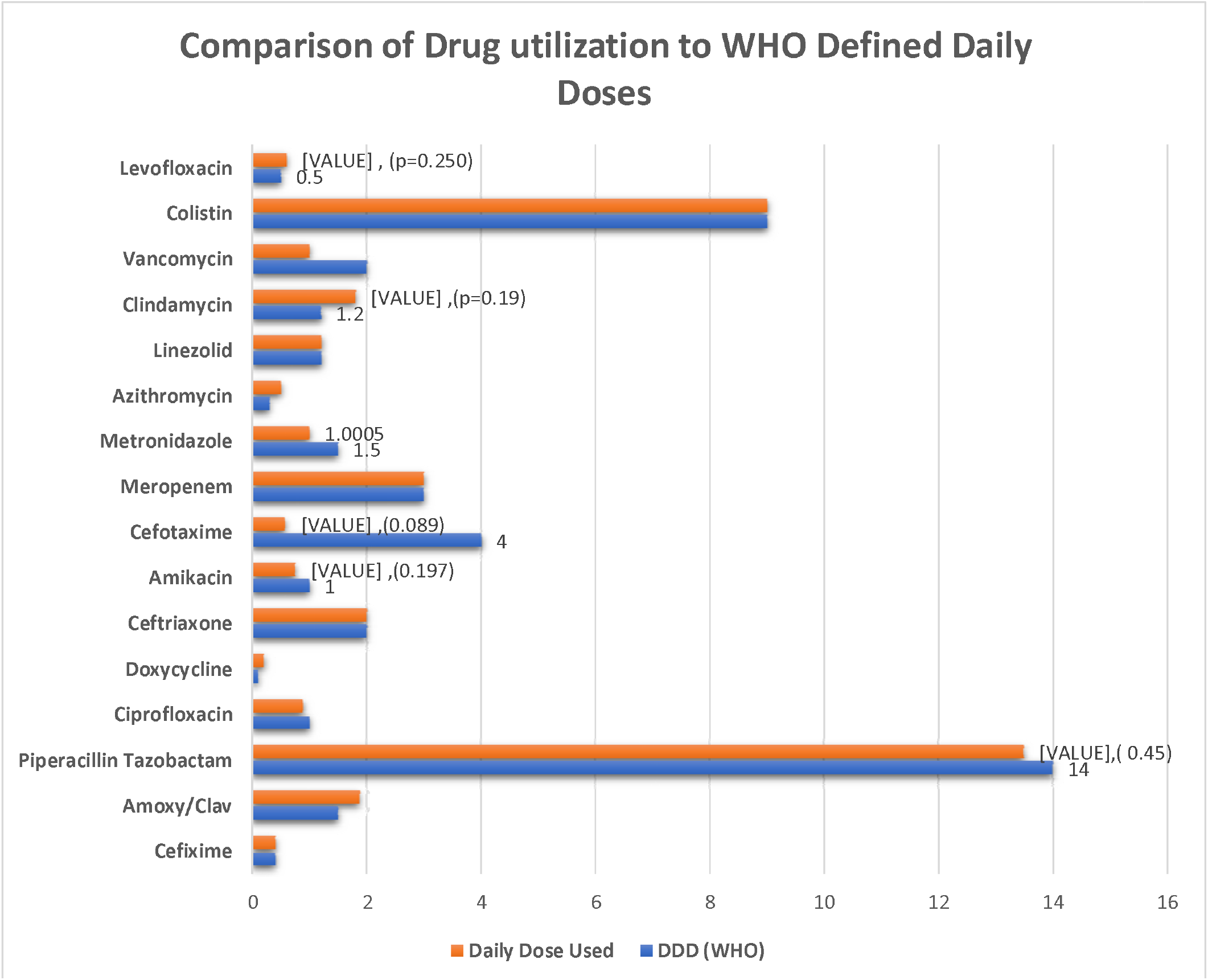
Comparison of Drug utilization to WHO Defined Daily Doses

Finally, the Mean Daily Drug Dose for prescribed drugs was compared with WHO defined DDD for each drug using a Student’s T test. The mean daily drug dose of Amoxy/Clav was significantly higher than the WHO DDD (2.28 vs. 1.50, p=0.014), while the mean daily drug dose of Metronidazole, Azithromycin, and Doxycycline were significantly lower than the WHO DDD (p<0.001, p=0.007, and p=0.008, respectively). The mean daily drug dose of Piperacillin / Tazobactam, Amikacin, Cefotaxime, Clindamycin, and Levofloxacin did not show significant differences compared to the WHO DDD (p>0.05).

## DISCUSSION

The findings of this study highlight several important issues related to antimicrobial prescribing practices and awareness among healthcare professionals. This study, conducted in a tertiary care institute in North India included prescriptions having at least one antibiotic in both inpatient (75.4%) and outpatient (24.6%) settings collected over a duration of 2 months by universal sampling. A total of 123 patient prescriptions were included which included 243 individual antibiotics. The mean number of antibiotics used per patient was 1.84 ± 0.83 which coincides with a study conducted in Brazil (8) with a mean of 2.4 antibiotics per patient. This finding is in line with the WHO prescribing indicators which state that each prescription should contain an average of 1.6–1.8 antibiotics (6). The use of multiple antibiotics can increase the risk of adverse effects, drug interactions, and development of antimicrobial resistance. The mean duration of antibiotics prescribed was 6.63 ± 3.83 days. A study conducted in a tertiary care hospital in India reported a mean duration of 5.7 ± 3.1 days (9), while another in Ethiopia reported a mean duration of 10.2 days (10). These variations could be attributed to differences in patient demographics, disease prevalence, prescribing habits, and hospital policies.

In the present study, prescriptions included 27 different antibiotics of which Metronidazole was the most prescribed antibiotic, which could be attributed to its broad-spectrum activity against anaerobic bacteria, which are commonly implicated in infections of the gastrointestinal tract, genitourinary tract, and skin. It is prescribed mostly in combination with amoxicillin in obstetrics and gynaecological care (mainly in post-delivery prophylaxis), and occasionally in caesarean section (11). Similar results were found study at Ghana Police Hospital (12). This was followed by Ceftriaxone, a third-generation cephalosporin from the Watch group. The high proportion of cephalosporins in the prescriptions is consistent with the WHO analysis of South-East Asian countries which also found a very high level of consumption of the same in all states in India (13).

The WHO AWaRe classification system was used to analyse the antibiotics prescribed in this study, and the results showed that most antibiotics fell under the Access (57.61%) category. These antibiotics have a narrow spectrum of activity, lower cost, a good safety profile and generally low resistance potential and should be widely available and affordable. While this is a positive finding, it is also important to note that a significant proportion of antibiotics prescribed were from the Watch category (38.27%), which includes antibiotics that are at risk of becoming ineffective due to overuse and misuse. The small percentage of antibiotics prescribed in the Reserve category (4.11%) is reassuring, as these antibiotics are reserved for use as a last resort and should only be used in highly specific circumstances. The overall prescribing of antibiotics in our study is lower than the WHO recommendations, which state that more than 60% of all prescribed antibiotics must be from the Access group by 2023. Figures in our study are consistent with another Indian study which had Access (53.31%), Watch (40.09%) from, and Reserve (3.40%) category (14), and study done in Zambia with (n=384) which had Access (55.5%), “Watch” (43.1%) and “Reserve” (1.4%) (6). Our findings differ from another study in India (15) in which Watch group antibiotics accounted for 53.19 % of the total antibiotics and a Bangladesh study in which 64.0% of the patients were treated with antibiotics from the Watch group, 35.6% were treated with antibiotics from the Access group, and only 0.1% were treated with antibiotics from the Reserve group. The higher proportion of Access category antibiotics prescribed in our study could be attributed to the fact that these antibiotics are commonly used for the treatment of common infections encountered in the hospital, such as urinary tract infections, respiratory tract infections, and skin and soft tissue infections. It is also important to note that our study was conducted in a resource-limited setting, where the availability of Watch and Reserve category antibiotics may be limited, leading to a higher use of Access category antibiotics. The low proportion of Reserve category antibiotics prescribed in our study is consistent with the recommended sparing use of these antibiotics by WHO and other guidelines, which recommend their use only as a last resort in the treatment of severe infections (16). In terms of departments and designations, our study found that the Medicine department prescribed the majority of antibiotics in both the Access and Watch categories, followed by the Surgery department.

This finding is consistent with previous studies conducted in India and other low- and middle-income countries, where the Medicine department was found to be the highest prescriber of antibiotics (17).

To assess knowledge and attitude of prescribing doctor about the utility of AWaRe a Questionnaire based survey was carried out. Although (73.1%) agreed that the emergence of antimicrobial resistance is inevitable the lack of awareness about the programmes and measures being taken to curb antimicrobial resistance is concerning. In contrast to this study, a study conducted in the United States found that healthcare professionals had a high level of knowledge about antimicrobial resistance and the appropriate use of antibiotics (18). This difference could be attributed to variations in healthcare systems and education programs in different countries. Moreover, in a study conducted in Germany, the majority of physicians agreed that the rational use of antibiotics is important for the prevention of antimicrobial resistance (19)

Despite the WHO AWaRe classification being included in the antibiotic policy of our institution only 33.3% of the responders knew about it which shines a light on the gap between the measures being taken and their implementation. The institute antibiotic policy authorises only the Senior Residents and Consultants to prescribe antibiotics from watch and reserve groups but in this study Junior Residents prescribed the highest number of antibiotics for both Access and Watch categories which is coincides with previous studies conducted in India and other countries. In a study conducted in a tertiary care hospital in India, junior residents prescribed the majority (68.7%) of antibiotics (17). The higher proportion of antibiotics prescribed by Junior Residents in our study could be attributed to their relatively higher workload and less clinical experience compared to Senior Residents and Consultants. Overall, the findings of this study suggest that although there is a moderate level of awareness among healthcare professionals about antimicrobial resistance, there is a need for further education and awareness programs to ensure the appropriate use of antibiotics in the hospital setting. It must be noted that most of the responders of the survey were also Junior Residents and hence education programmes can hence play a vital role in improving the results by working at grassroot levels.

Another analysis of the study depicts that there is a significant difference between the WHO DDD and the prescribed daily dose in the analysed prescriptions. Several studies conducted in the West have compared the mean daily drug dose of prescribed drugs with the WHO defined DDD. In a study conducted in a hospital in Italy found that the mean daily dose of antibiotics was generally lower than the WHO DDD for most antibiotics (20). These findings are similar to the present study, which found higher mean daily doses of amoxicillin/clavulanic acid and piperacillin/tazobactam, and a lower mean daily dose of azithromycin and levofloxacin. The WHO DDD is a standardized measure of drug consumption used to compare the drug consumption between different countries and regions. It represents the assumed average maintenance dose per day for a drug used for its main indication in adults. On the other hand, the prescribed daily dose refers to the actual dose that a physician prescribes to a patient.

The difference between the WHO DDD and the prescribed daily dose can have several implications. Firstly, it can lead to overuse or underuse of medications, which can affect the therapeutic outcomes of patients. For example, if the prescribed daily dose is lower than the WHO DDD, the patient may not receive the optimal therapeutic effect of the medication. Conversely, if the prescribed daily dose is higher than the WHO DDD, the patient may be at risk of adverse effects or toxicity. Secondly, the difference between the WHO DDD and the prescribed daily dose can affect the comparability of drug consumption data between different regions and countries. If different regions or countries use different prescribed daily doses, it can be difficult to compare their drug consumption patterns. Therefore, adherence to the WHO DDD is important to ensure that drug consumption data are standardized and comparable across different regions and countries. In conclusion, the difference between the WHO DDD and the prescribed daily dose is an important issue that needs to be addressed in the prescribing practices of physicians. Adherence to the WHO DDD can help to ensure optimal therapeutic outcomes for patients and comparability of drug consumption data between different regions and countries.

Overall, the findings of this study emphasize the importance of improving antimicrobial prescribing practices and increasing awareness among healthcare professionals regarding the WHO AWaRe classification system and the threat of antimicrobial resistance. Effective antimicrobial stewardship programs that promote appropriate antibiotic use can help reduce the risk of antimicrobial resistance and improve patient outcomes. Future research should focus on implementing such programs in hospital settings and evaluating their effectiveness in reducing inappropriate prescribing practices.

## Data Availability

All data produced in the present study are available upon reasonable request to the authors

## Notes

### Competing Interest Statement

The authors have declared no competing interest.

### Funding Statement

This study did not receive any funding

### Author Declarations

The study was conducted after approval by the Institutional Ethics Committee (All India Institute of Medical Sciences, Rishikesh, India)

